# Wisely Frugal: Ensuring sustainable funding for novel cancer therapeutics using cost-effectiveness thresholds in resource limited settings

**DOI:** 10.1101/2023.02.17.23285787

**Authors:** Nuradh Joseph, Vimukthini Peiris, Vodathi Bamunuarachchi, Prasad Abeysinghe, Nadarajah Jeyakumaran, Devinda Jayathilake, Kanthi Perera, Rohini Fernandopulle, Sanjeeva Gunasekera

**Affiliations:** District General Hospital - Hambantota, Sri Lanka; Sri Lanka Cancer Research Group, Maharagama, Sri Lanka; District General Hospital - Vavuniya, Sri Lanka; Apeksha Hospital, Maharagama, Sri Lanka; Sir John Kotalawela Defence University, Kandawala, Sri Lanka

**Keywords:** Essential drugs, medical oncology, cost-effectiveness, lower middle income countries, novel drugs, cancer therapeutics

## Abstract

**Introduction:** Cancer care in Sri Lanka is predominantly provided through its public funded state health system which is free at the point of delivery. The health system faced unprecedented funding restrictions brought about by the post-pandemic recession. We performed a cost-effectiveness analysis of novel cancer drugs with a view to prioritising novel cancer therapeutics to its sustainability during these challenging times.

**Methods:** The direct cost of drug procurement was obtained, and the cost per life year gained was computed for each indication. Two thresholds - per capita GDP per life year gained (GDPx1) and three times per capita GDP per life year gained (GDPx3) were considered to determine cost effectiveness. The cumulative annual cost of these treatments were then determined by multiplying the cost per treatment course per patient by the estimated number of treated patients per year for each indication.

**Results:** Data obtained on 42 novel cancer drugs spanning across 90 indications were included in the analysis. The cumulative annual treatment cost when the threshold was set at GDPx1 was US$ 6 million and it increased to US$ 16.3 million if the threshold was expanded GDPx3. Only 28 indications met the GDPx3 threshold while there were 18 drugs that did not meet the thresholds for any indication. Without a threshold, if every eligible patient were to receive the indicated currently used novel drugs, the total cost of treatment would reach almost US$ 300 million per year.

**Conclusion:** Cost-effectiveness thresholds will lead to considerable savings and help prioritise procurement and supply of cost-effective novel agents in the state health system in Sri Lanka.

**Advances in Knowledge:** In this work, we show that significant savings can be achieved by performing simple cost-effectiveness analyses and defining thresholds. The absence of robust quality of life and costing data should not deter policy makers from such conducting analyses from available information.

## Introduction

Sri Lanka has an age adjusted annual incidence of 129 cancer cases per 100,000 population with nearly 32,000 new patients being diagnosed each year, according to data from the national cancer registry^1^. However, the actual incidence might be considerably higher due to under-reporting of cases^2^.

Cancer care in Sri Lanka is predominantly provided through its public funded state health system which is free at the point of delivery^2,3^. Funded by general taxation, the state health system functions as a network of primary,secondary and tertiary care hospitals under the administrative control of the Ministry of Health^2,3^. Clinical Oncology services are provided by 26 cancer centres located throughout the island^2,3^.

Each year, the Ministry of Health of the government of Sri Lanka allocates around 210 million US$ for the procurement of drugs for hospitals under its purview^4^. In the beginning of 2022, Sri Lanka faced a foreign exchange crisis arising from years of imprudent macroeconomic fiscal policies aggravated by the Covid-19 pandemic^5^.

The crisis was devastating and almost led to a calamitous breakdown of Health services and procurement of pharmaceuticals imported from abroad proved extremely challenging^5^. Since virtually all oncology drugs are imported, the economic crisis posed a major threat to cancer care.

We performed a cost-effectiveness analysis of novel cancer drugs with a view to compiling a list of essential drugs and approved indications to help prioritise procurement and help mitigate the impact on cancer care.

## Methods

### Computing cost of drugs

The cost of each individual drug procured during the year 2021 was obtained from the Medical Supplies division of the Ministry of Health published on its website^6^. This was converted to United States Dollars based on the average exchange rate for the year 2021.

The direct drug cost for a standard course of treatment was computed as for an adult male weighing 50kg with a body surface area of 1.3. Indirect costs as well as costs of administration such intravenous cannulas, infusion sets etc. were not considered.

Since the focus is on novel therapeutics, conventional anti-neoplastic agents where the total cost of treatment was less than 1000 US$ were excluded from the analysis.

If the novel therapeutic agent was not an additive treatment the costs of the drugs used in the comparator arm was subtracted from the cost of the novel agents.

### Computing Survival gain

Since data on quality adjusted life years was not available in the local setting we considered life years gained as the outcome parameter. This was obtained from publications of pivotal randomised controlled trials for drug and indication.

In the palliative setting median overall survival was the outcome variable. In trials, where there was significant crossover of treatment, progression free survival gain was considered, especially if reliable estimates of overall survival were not available.

In the curative setting life years gained per patient was computed using the following method. It was assumed that a survivor at the end of the specified follow-up period would have a life expectancy close to that of the normal population. Since the life expectancy at birth of Sri Lankans is currently 77 years we chose 75 years for this assumption. The number of life years gained by a survivor following completion of follow-up was computed by subtracting from 75 years the sum of median age at enrollment and DFS time interval, and multiplying this value by the absolute DFS gained. The life years gained during the trial follow-up duration was computed by dividing the DFS time interval by two and multiplying this value by the absolute DFS gain. Total life years gained was taken as the sum of life years gained during and after follow-up.

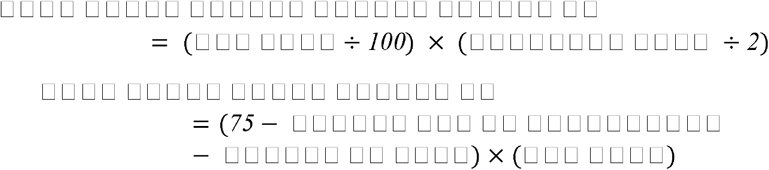

### Cost per life year gained

The cost per life year gained was then computed by dividing the total cost of treatment by life years gained by the treatment.

### Trials of treatment de-escalation

We also considered studies of statistically proven equivalence or non-inferiority of de-escalated treatment. We assumed that the survival gains of the de-escalated treatment was the same as standard treatment and the cost of de-escalated treatment was computed and divided by life-years gained of the standard treatment to determine cost per life year gained by the de-escalated treatment.

### Cost-effectiveness thresholds

Three thresholds based on World Health Organization recommendations were used for this analysis - viz: less than the per capita annual gross domestic product (GDP) per life year gained (GDP x 1; highly cost-effective), 1-3 times the per-capita annual GDP per life year gained (GDP x 3; cost-effective) and GDP 3-4 (GDP x 4; potentially cost-effective with price reduction) were considered. The per capita GDP of Sri Lanka for the year 2021 was obtained to compute these thresholds ^11,12^.

### Total Cost of Treatment

The total cost per year for the entire country was computed by multiplying the total cost per patient by the estimated number of treated patients based on national incidence data, and proportion of patients likely to receive treatment for each indication. These estimates were made by three oncologists independently and the mean value was taken for the analysis.

The list of references used to obtain data for each treatment is listed in supplementary appendix.

## Results

The average exchange rate for the year 2021 was 200 Sri Lankan Rupees per United States Dollar (US$)^7^. The per capita GDP for the year 2021 was US$ 3815. The cost of treatment per standard treatment course was obtained for 83 oncology drugs^8^. Conventional agents which were excluded from the analysis are shown in Supplementary Table S1 along with the cost per standard treatment course.

After exclusion of these drugs, 41 drugs spanning across 83 indications in the palliative setting and 6 drugs across 10 indications in the adjuvant setting were included in the analysis. A full description of the analysis including the clinical trials on which outcome data was obtained from is included in the supplementary appendix.

There were two drugs for which de-escalated treatment was of relevance. When analysing abiraterone, we considered a low dose treatment regimen of 250 mg with food and the standard dose of 1000mg on an empty stomach for each indication. Similarly for adjuvant trastuzumab we considered 6 months of treatment as well as 12 months of treatment.

Tables 1 and 2 list the highly cost-effective and cost-effective drug indications respectively along with the total annual cost of procurement for these drugs. Supplementary Tables S2 and S3 list the drugs and indications which are potentially cost-effective and not cost-effective respectively. Figure 1 shows the plot for cumulative annual procurement cost against per capita GDP per life year gained, while Figure 2 shows the same plot with the thresholds limited up to four times the per capita GDP per life year gained.

**Table 1.**
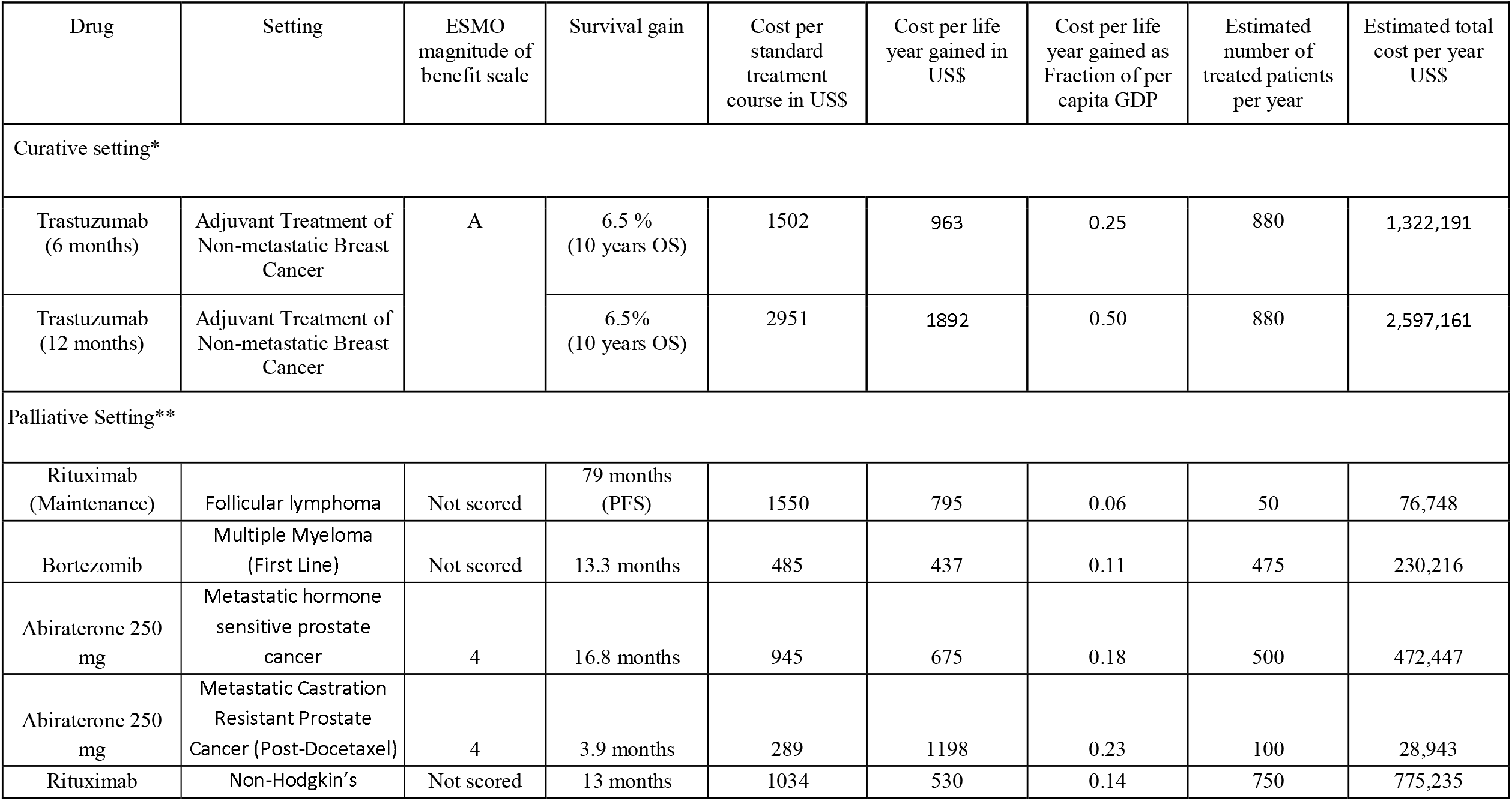
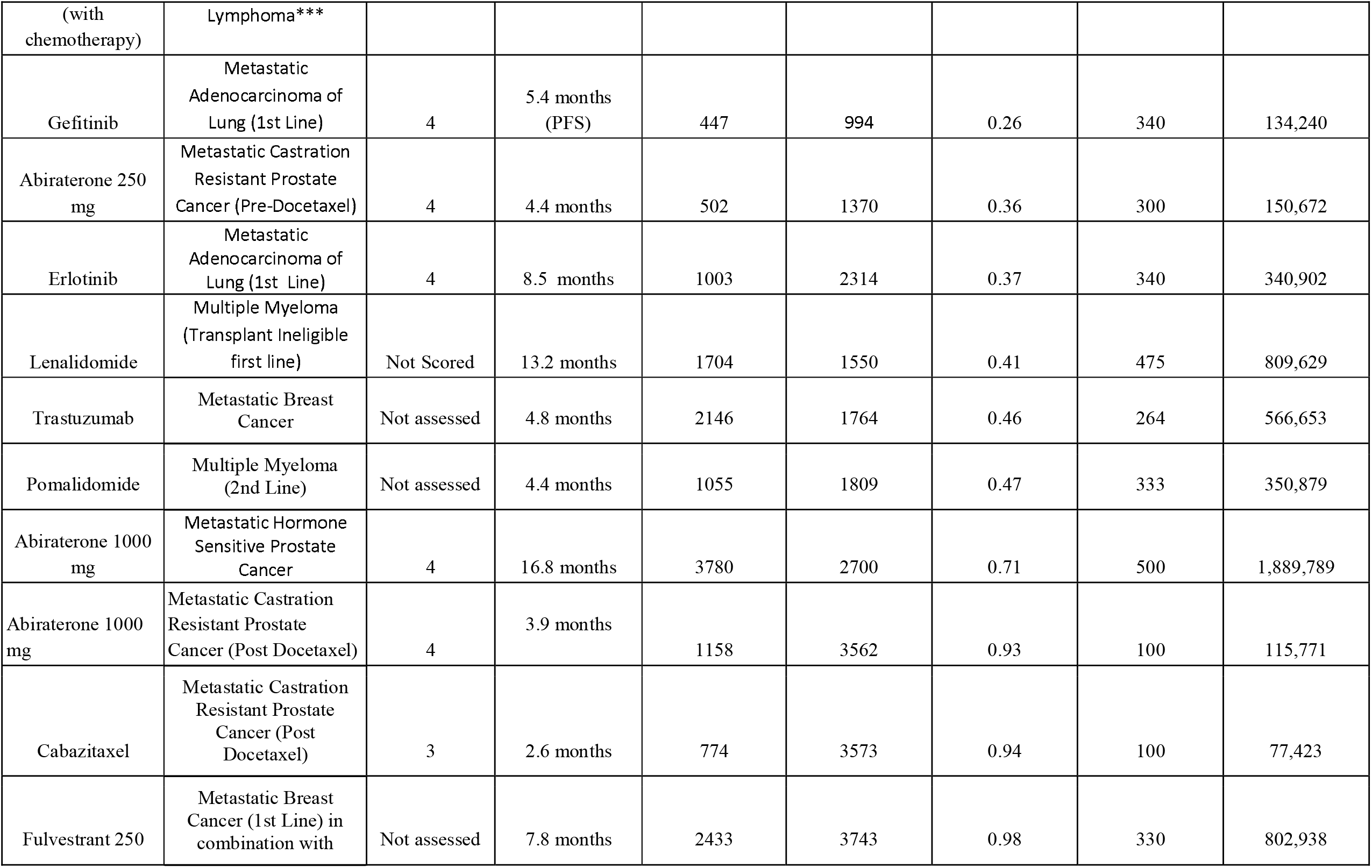
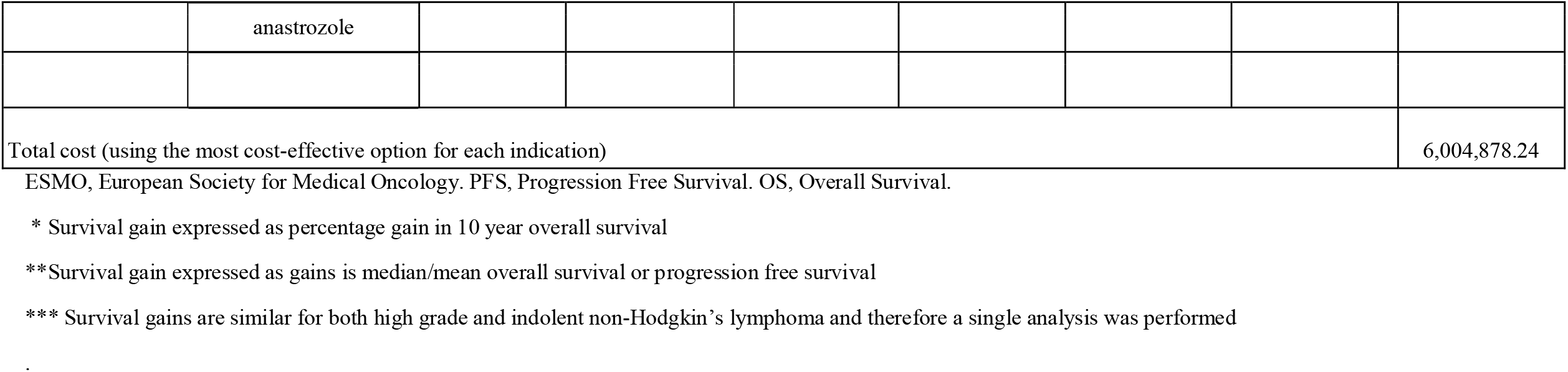
Highly cost-effective treatments (Cost per life year gained less than per capita GDP)

**Table 2.**
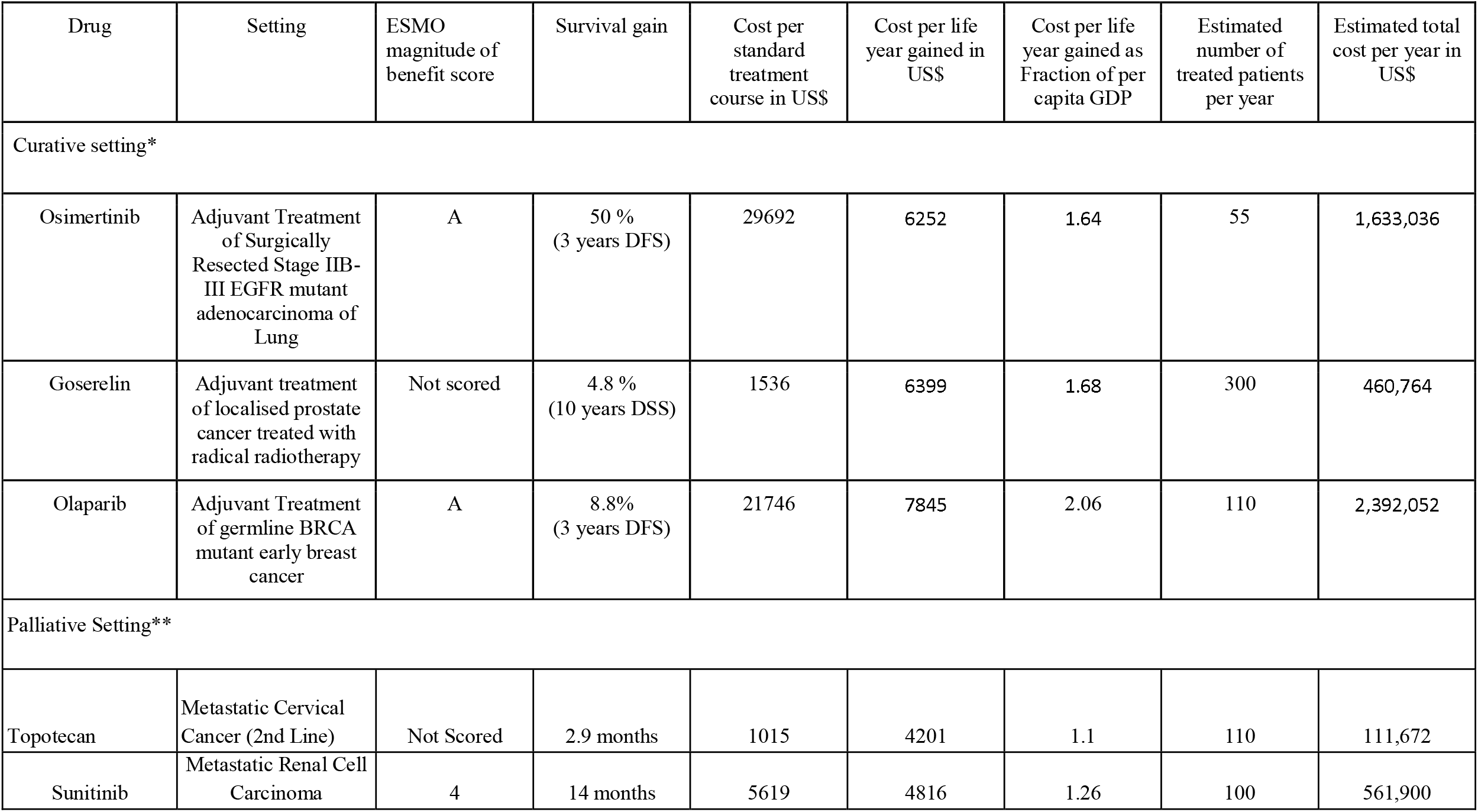
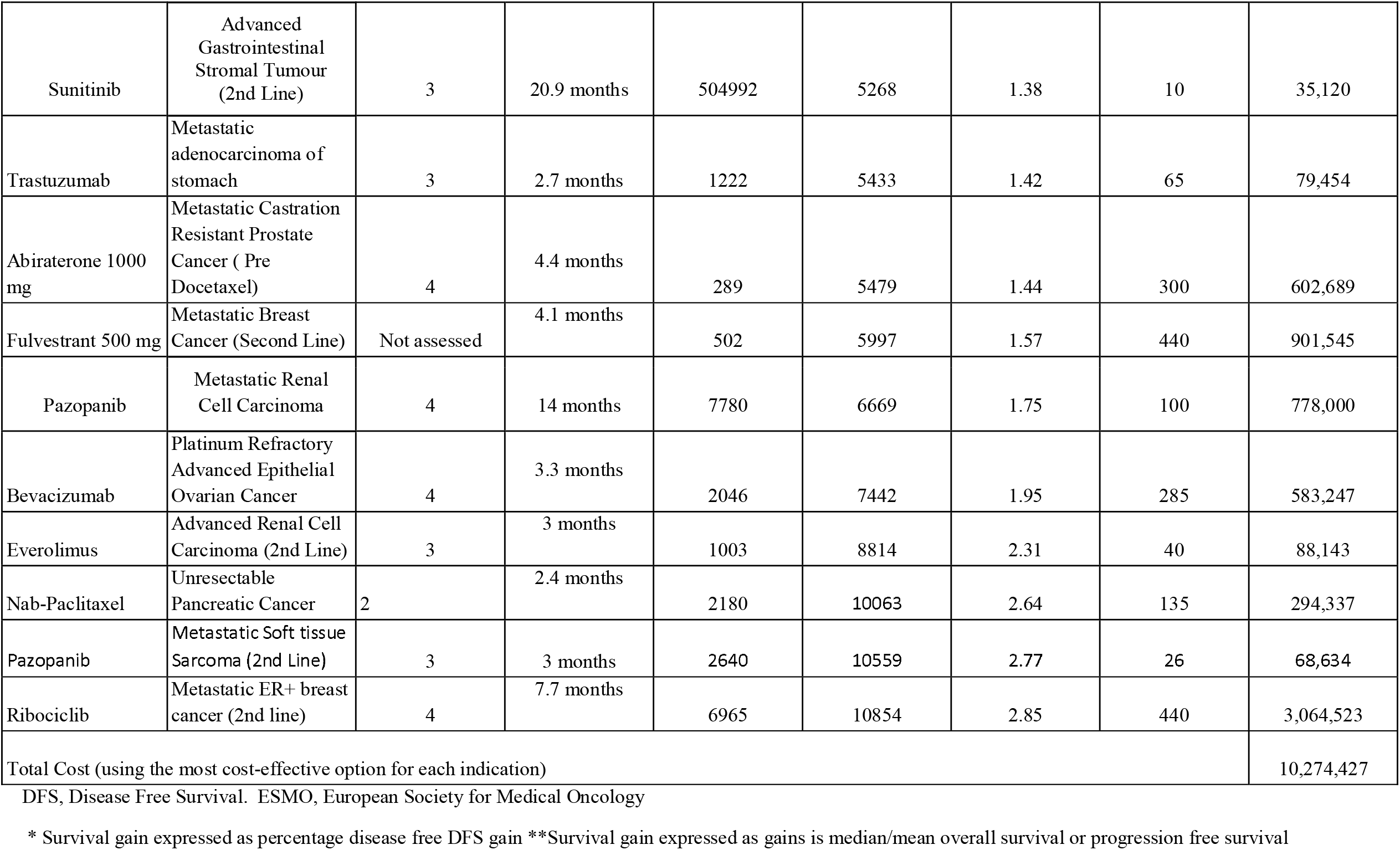
Cost-effective treatments (drug cost per life year gained between 1-3 times per capita GDP)

**Figure 1.**
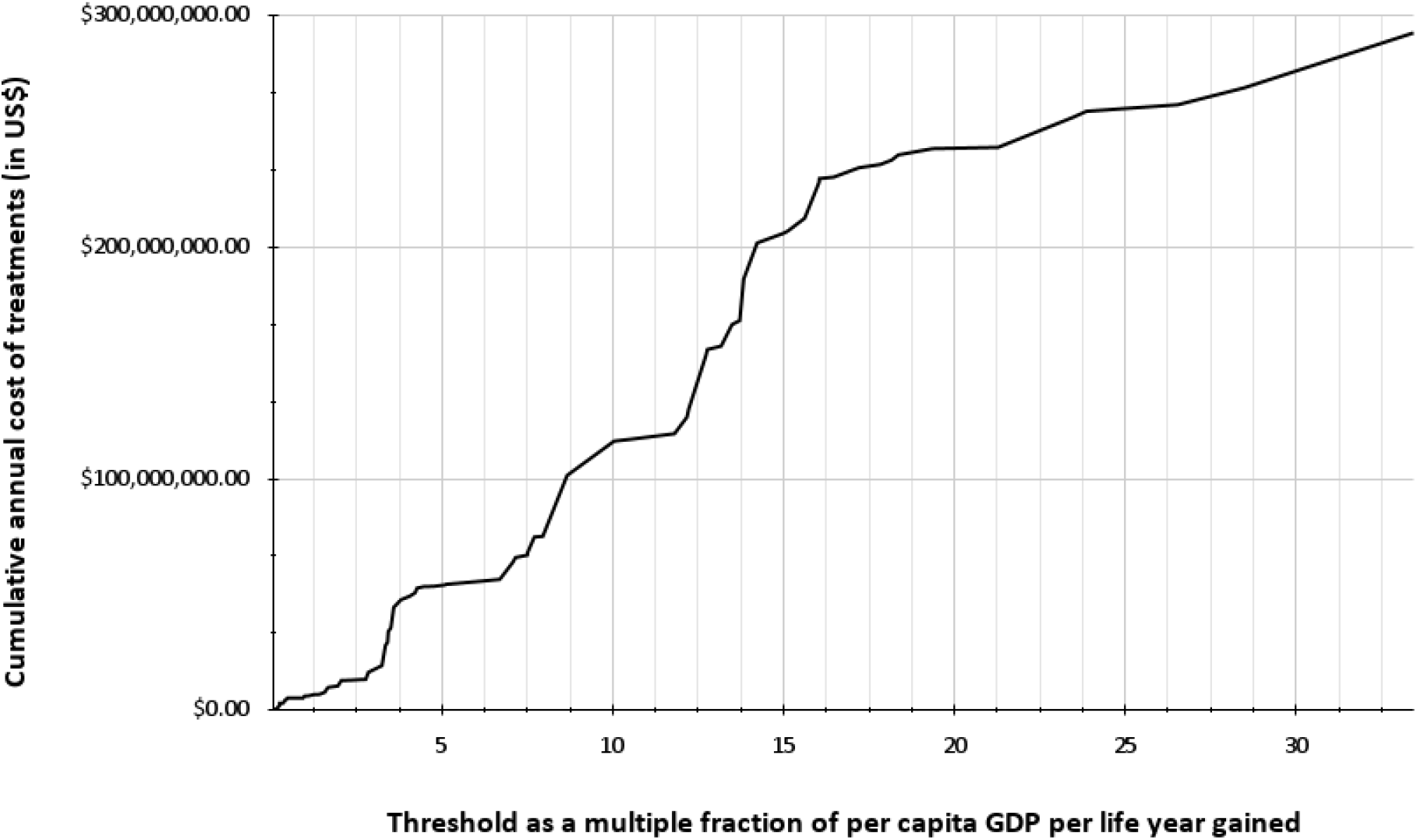
Cumulative cost of treatments of novel drugs in relation to cost-effectiveness thresholds based on per capita GDP per life year gained.

**Figure 2.**
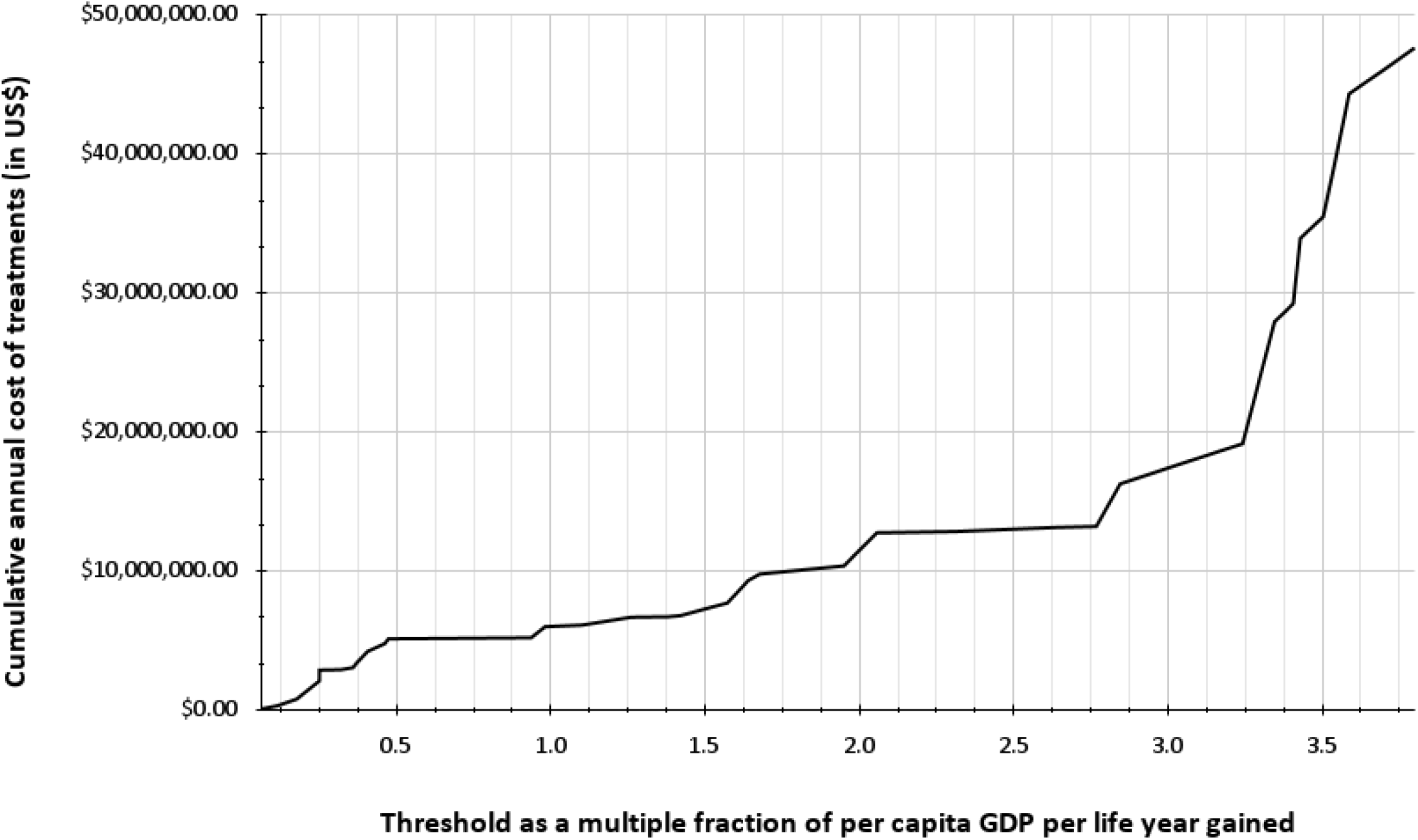
Cumulative cost of treatments of novel drugs in relation to cost-effectiveness thresholds upto per capita GDP times four per life year gained.

The total cost of treatment is US$ 6 million if a threshold of per capita GDP per life year gained (GDP x 1) was set and US$ 16.3 million if it was per capita GDP x 3 per life year gained. If the threshold is increased to GDP x 4, the total cost would rise to US$ 47.3 million. If no threshold was set, Sri Lanka’s health system would need US$ 295 million to fund these novel drugs.

## Discussion

The allocation of funds for healthcare in a public funded state health system is determined by political authorities and is often influenced by the general macroeconomic situation of the country^9^. Once the allocation is decided, it is imperative that cost-effectiveness based thresholds be considered to ensure maximum benefit from the drugs procured by each health system^10^.

In this work we present a rational basis for prioritising procurement of supply of oncology drugs in Sri Lanka which we believe would find resonance with health care systems of other low and middle income countries (LMICs). We also provide further evidence for the validity of the cut-off of GDP x 3 per life year gained as the threshold for determining cost-effectiveness ^11,12^.

Based on this threshold the budget for procurement of novel cancer therapeutics would be approximately 16.8 million US$ which would be around 9% of the total drug budget of the state health system. Increasing the threshold to GDP x 4 would nearly triple the amount of funding required, further validating the robustness of the WHO threshold of GDP x 3. Without cost-thresholds, the cumulative annual cost of the currently procured novel drugs would be nearly US$ 300 million assuming every eligible patient would receive treatment. This is around 1.5 times the total annual budget of all drugs in the state health sector. In the absence of well defined cost-effectiveness thresholds, a health system such as ours will be open to frequent shortages of essential drugs when the budgeted allocation is expended or if unforeseen factors result in a foreign exchange crunch.

In a time of financial crisis, the lower threshold of per capita GDP per life year gained (GDP x 1) can be used to prioritise procurement. As shown by our data, a total allocation of approximately US$ 6 million would be sufficient to ensure supply of these highly cost-effective drugs. Furthermore, defining a cost-effectiveness threshold would also provide an incentive for pharmaceutical suppliers to reduce the price of drugs thereby leading to cost savings. These savings could be channelled to more cost-effective treatment modalities such as radiotherapy and surgery. Indeed, studies have shown that the dearth of quality radiotherapy resources in Sri Lanka has adversely impacted on outcomes of potentially curative cancers^13,14^. Investing in screening and streamlining early detection pathways may also lead to significant improvements in survival ^15,16,^.

Except for ibrutinib which is not registered in Sri Lanka, all treatments mentioned in the World Health Organisation (WHO) essential drugs list were found to be cost-effective in our setting as well^17^. However, there were several other treatments that were not included in the WHO list that were found to be cost-effective in our study, which are listed in Box 1. Two such treatments, adjuvant osimertinib in resected in adenocarcinoma of the lung and adjuvant olaparib in early germline BRCA mutation positive breast cancer, are very recent developments ^18,19^. Nevertheless, this underscores the importance of performing local cost-effectiveness assessments to take into account cost variations in different health systems.

Another salient finding of our work is the cost-savings that can be achieved by using lower doses of abiraterone for metastatic prostate cancer and a shorter duration of adjuvant treatment with trastuzumab in early breast cancer, both of which have robust evidence in the form of non-inferiority randomised clinical trials^20,21^.

For abiraterone a lower dose of 250mg with food was shown to have equal efficacy in terms of biochemical response in castration resistant prostate cancer^20^. The clinical equipoise can be safely extrapolated to the hormone sensitive phase of the disease as well and this is borne out by the inclusion of the lower dose option in the NCCN guidelines for both settings^22^.

Even though the landmark PERSEPHONE trial of more than 4000 patients proved non-inferiority for 6 months of adjuvant trastuzumab with 12 months of treatment, the oncology community has been slow to adopt this partly due to concerns with its subgroup analysis showing superiority of 12 months of treatment in patients receiving concurrent trastuzumab with chemotherapy^23,^. However clinicians in LMICs such as ours would be well advised to opt for six months of adjuvant trastuzumab due to its substantial cost savings and the uncertain benefit of one year of adjuvant treatment to one year, which if at all, is likely to be marginal. The newer anti HER-2 monoclonal antibody Pertuzumab was not even remotely cost-effective either in the adjuvant or metastatic setting. Trastuzumab emtansine has not been used in the state health sector and we were therefore unable to perform an analysis of cost-effectiveness. However, unless very substantial price reductions are made these agents are unlikely to be cost-effective in our setting.

With regard to multiple drugs for the same indication, we found that abiraterone was substantially more cost-effective than enzalutamide in metastatic prostatic cancer both in its hormone sensitive and castration resistant phases. Similarly gefitinib and erlotinib were superior to osimertinib in the first line treatment of metastatic adenocarcinoma of the lung, while pembrolizumab was more cost-effective than nivolumab in metastatic melanoma.

Despite gaining approval for multiple malignancies, the immunotherapeutic agents Pembrolizumab and Nivolumab failed to reach the cost-threshold in almost all indications with the exception of metastatic melanoma, where it was potentially cost-effective.

It is unlikely that these drugs would be affordable in health systems of LMICs such as ours in the near future. However, encouraging results from recent trials exploring the efficacy of lower doses of these agents. provides some hope, and more studies in this space are an imperative need ^24^.

Analysis of agents used in the first line treatment of Chronic Myeloid Leukaemia posed many issues since it is takes the form of a chronic disease entity where treatment extends beyond 10 years, When considering imatinib in the first line treatment of CML, its cost is cheaper than the comparator interferon-alpha and low dose cytarabine. Since the annual treatment cost per patient for imatinib was only 180 US$ we excluded it from this analysis since its cost-effectiveness is evident. Due to short follow-up in the clinical trials of first line treatment of CML, with the novel tyrosine kinase agents nilotinib and dasatinib, it was not possible to determine the survival gain accurately ^25^. However the overall survival gain over imatinib is likely to be very modest in the first line setting and these agents are substantially more expensive than imatinib^25^. Over a 10-year period nilotinib and dasatinib would cost US$ 53000 and 44347 more than imatinib, respectively.

For our analysis we only considered the direct cost of the drug. The cost of drug administration, investigations, staff costs etc. were not included. Furthermore, since data on quality of life was not available, we were compelled to consider life years gained as the outcome variable rather than quality adjusted life years. The paucity of data also meant that more robust cost-effectiveness analysis methods such as a Markov model could not be performed. As such, our data is likely to overestimate the benefit of treatment. More studies on evaluating quality of life and cost of treatment in the state health sector are required as a matter of urgency.

The inability to determine quality adjusted life year gains for novel therapeutics should not deter health systems such as ours from using more simple metrics when setting cost-effectiveness thresholds. As shown by our results even rudimentary cost-effectiveness analysis could provide valuable insights when making decisions on funding. Some data is better than none in this regard.

Our intention was to highlight the importance of cost-effectiveness thresholds in determining novel cancer drug procurement and usage in health systems such as ours. We believe this has been achieved by our work, notwithstanding the limitations mentioned above.

## Supporting information

Reference List

## Data Availability

All data produced in the present study are available upon reasonable request to the authors

## List of Tables and Figures

**Supplementary Table S1.**
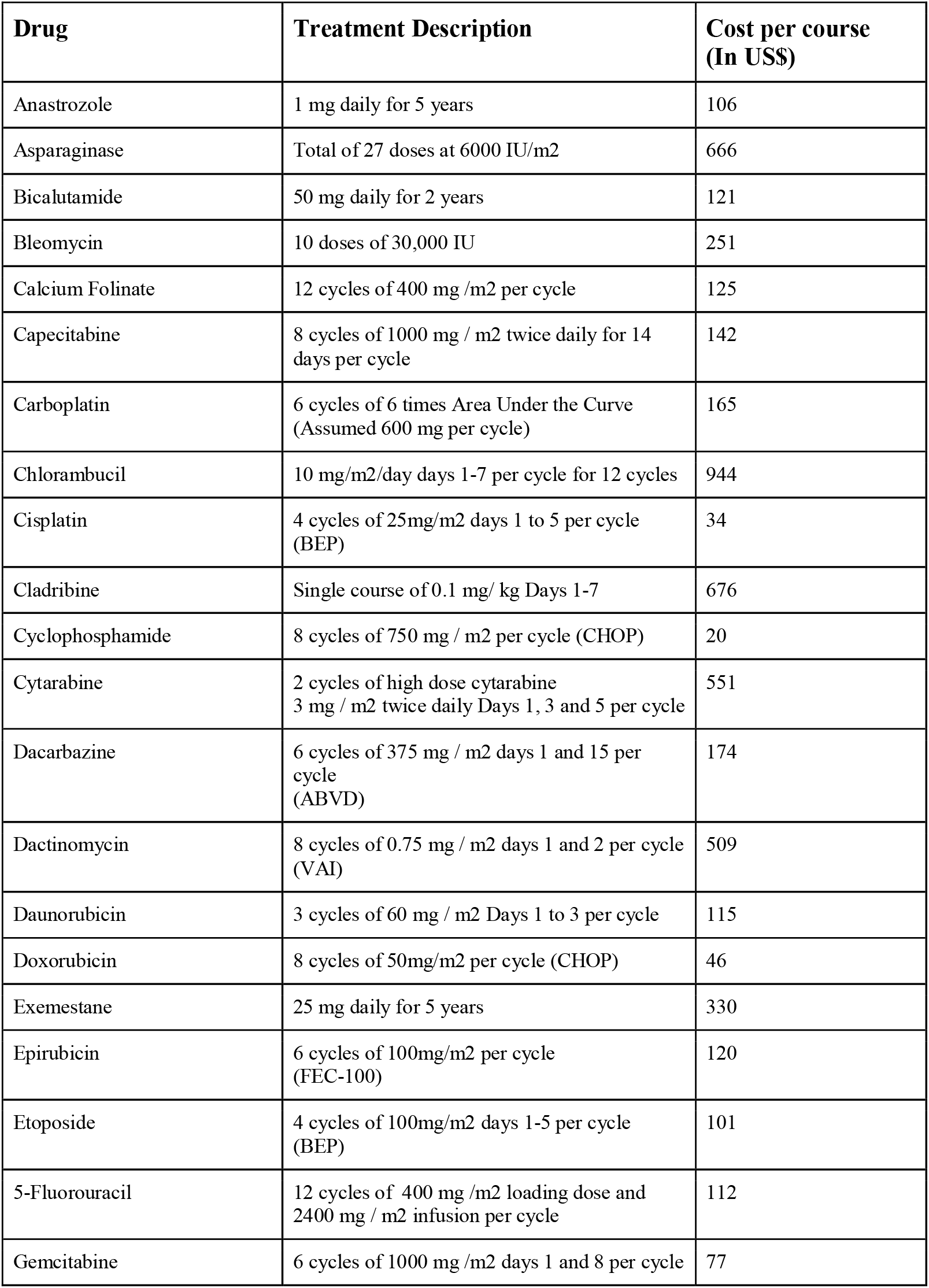
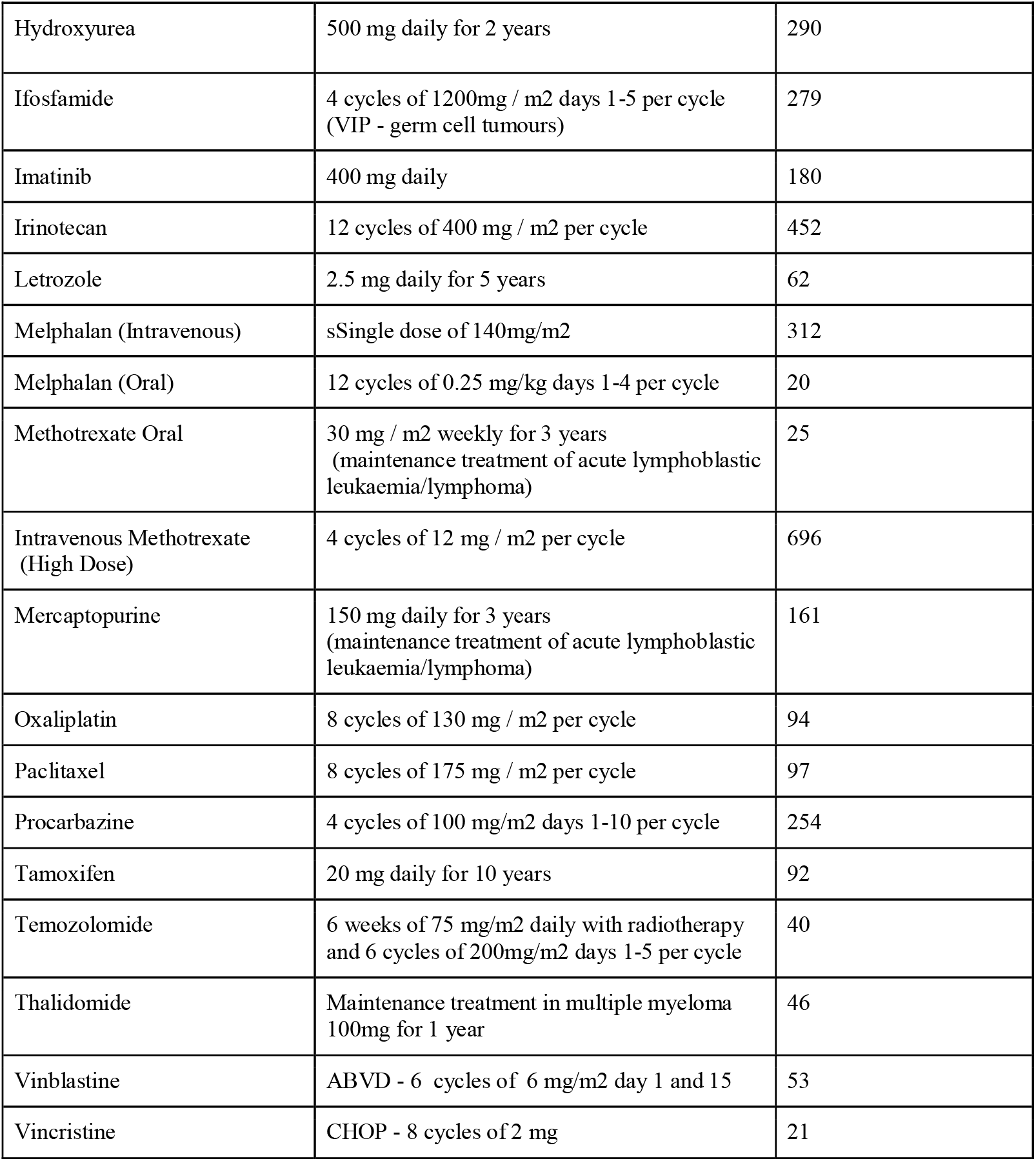
Conventional agents excluded from the analysis and cost per treatment course.

**Supplementary Table S2.** Potentially Cost-effective treatments (Drug cost per life year gained between 3-4 times per capita GDP)

| Drug | Setting | Survival gain<br>(ESMO<br>magnitude of<br>benefit score in<br>parenthesis) | Cost per standard<br>treatment course<br>in US\$ | Cost per life year<br>gained in US\$ | Cost per life year<br>gained as<br>Fraction of per<br>capita GDP | Estimated number<br>of treated patients<br>per year | Estimated total<br>cost per year in<br>US\$ |
| --- | --- | --- | --- | --- | --- | --- | --- |
| Curative setting* |  |  |  |  |  |  |  |
| Pembrolizumab | Adjuvant Treatment of<br>Surgically Resected Stage<br>III melanoma | 18.4 % DFS<br>(A) | 59799 | 13684 | 3.59 | 100 | 5,979,899 |
| Palliative Setting*** |  |  |  |  |  |  |  |
| Olaparib | Maintenance treatment of<br>newly diagnosed BRCA<br>mutant Epithelial Ovarian<br>Cancer | 42.2 months<br>(4) | 43492 | 12367 | 3.24 | 67 | 2,892,209 |
| Ribociclib | Metastatic hormone<br>sensitive breast cancer (1st<br>lin) | 12.5 months<br>(4) | 13296 | 12765 | 3.35 | 440 | 8,775,678 |
| Sorafenib | Advanced Hepatocellular<br>Cancer | 2.8 months<br>(3) | 3010 | 12898 | 3.38 | 250 | 752,412 |
| Bevacizumab | Metastatic Colorectal<br>Cancer (2nd Line) | 2.1 months<br>(3) | 2274 | 12994 | 3.41 | 250 | 568,467 |
| Nivolumab | Metastatic Melanoma | 26.1 months<br>(4) | 28342 | 13031 | 3.42 | 165 | 4,676,382 |
| Bevacizumab | Advanced Ovarian Cancer<br>(2nd Line treatment of<br>Platinum Sensitive Relapse) | 4.9 months | 5457 | 13365 | 3.5 | 285 | 1,555,327 |
| Osimertinib | EGFR mutant<br>adenocarcinoma of lung<br>(2nd Line) | 10 months<br>(4) | 11199 | 13438 | 3.52 | 255 | 2,855,677 |
| Bevacizumab | Stage IV ovarian epithelial<br>carcinoma | 4.8 months | 5798 | 14496 | 3.8 | 650 | 3,305.069 |
| Total cost |  |  |  |  |  |  | 31,368,035 |

DFS, Disease Free Survival. ESMO, European Society for Medical Oncology
\* Survival gain expressed as percentage disease free DFS
\*\* See Supplementary Table S1 for explanation on calculation of DFS gain.
\*\*\*Survival gain expressed as gains is median/mean overall survival or progression free survival

**Supplementary Table S3.** Treatments which are not cost-effective (Cost per life year gained more than 4 times per capita GDP)

| Drug | Indication | Cost per life year gained<br>(in multiples of per<br>capita GDP) | Estimated total cost per<br>year |
| --- | --- | --- | --- |
| Palbociclib | Hormone Sensitive Metastatic Breast Cancer 2nd Line | 4.03 | \$3,042,792 |
| Bevacizumab | Metastatic Cervical Cancer | 4.06 | \$1,575,791 |
| Pembrolizumab | Metastatic Melanoma | 4.10 | \$4,062,814 |
| Lapatinib | Metastatic HER2 positive Breast Cancer 2nd Line | 4.19 | \$1,171,205 |
| Decitabine | Acute Myeloid Leukaemia | 4.21 | \$286,479 |
| Palbociclib | Metastatic Breast CA 1st line | 4.22 | \$9,128,377 |
| Nilotinib | Imatinib resistant Chronic Myeloid Leukaemia | 4.27 | \$1,980,848 |
| Nivolumab | Adjuvant treatment of Stage III melanoma | 4.47 | \$11,053,266 |
| Ribociclib | Pre-menopausal Hormone Sensitive Metastatic Breast Cancer 1st Line | 4.47 | \$668,623 |
| Trabectedin | Metastatic Soft tissue Sarcoma 2nd line | 4.75 | \$105,986 |
| Azacitidine | Acute Myeloid Leukaemia | 5.08 | \$604,654 |
| Regorafenib | Metastatic Treatment Refractory Colorectal Cancer | 5.12 | \$284,925 |
| topotecan | Platinum Refractory Advanced Ovarian Cancer | 6.05 | \$1,205,545 |
| Ceritinib | Metastatic ALK+ Non-small cell lung cancer | 6.69 | \$922,244 |
| Osimertinib | EGFR mutation metastatic adenocarcinoma of lung 1st Line | 7.08 | \$7,462,746 |
| Bevacizumab | Metastatic Colorectal Cancer First Line | 7.15 | \$1,989,636 |
| Enzalutamide | Metastatic Castration Resistant Prostate Cancer Post-Docetaxel | 7.50 | \$1,058,314 |
| Bevacizumab | Metastatic non-squamous non-small cell lung cancer | 7.51 | \$1,217,657 |
| Pembrolizumab | Metastatic non-small cell lung cancer PD-L1>50% First Line single agent use | 7.70 | \$6,577,889 |
| Crizotinib | Metastatic ALK+ Non-small cell lung cancer | 7.95 | \$335,216 |
| Pembrolizumab | Adjuvant treatment of triple negative breast cancer | 8.66 | \$26,311,558 |
| Olaparib | BRCA mutant ovarian cancer maintenance treatment | 10.04 | \$14,823,277 |
| Pembrolizumab | Unresectable squamous cell carcinoma of head and neck PD-L1 positive single agent use | 10.54 | \$844,221 |
| Dasatinib | Imatinib resistant Chronic Myeloid Leukaemia | 11.80 | \$2,359,852 |
| Pembrolizumab Adjuvant RCC | Adjuvant treatment of renal cell carcinoma | 12.17 | \$7,175,879 |
| Lenvatinib | Radioiodine refractory advanced thyroid cancer | 12.22 | \$3,515,879 |
| Pembrolizumab | Metastatic non-squamous non-small lung cancer in combination with chemotherapy (First Line use) | 12.73 | \$23,321,608 |
| Cetuximab | Metastatic Treatment Refractory Colorectal Cancer RAS wild type single agent use | 12.77 | \$2,384,422 |
| Pembrolizumab | Metastatic PD-L1 positive non-small cell lung cancer (second line single agent use) | 13.17 | \$1,494,975 |
| Panitumumab | Metastatic Colorectal Cancer RAS wild type First Line use with chemotherapy | 13.49 | \$9,327,889 |
| Nivolumab | Unresectable squamous cell carcinoma of head and neck second line | 13.72 | \$1,700,503 |
| Pembrolizumab | Metastatic Squamous cell carcinoma of lung first line use in combination with chemotherapy | 13.83 | \$17,939,698 |
| Pertuzumab | Metastatic HER2 positive Breast Cancer 1st Line | 14.22 | \$15,619,503 |
| Pembrolizumab | Unresectable PD-L1 positive squamous cell carcinoma of head and neck in combination with chemotherapy | 15.00 | \$4,221,106 |
| Olaparib | BRCA mutant platinum sensitive ovarian cancer single agent use (third line) | 15.13 | \$959,353 |
| Enzalutamide | Metastatic Castration Resistant Prostate Cancer Pre-Docetaxel | 15.62 | \$5,507,162 |
| Pembrolizumab | Metastatic Triple Negative Breast Cancer PD-L1 10% or more | 16.04 | \$15,477,387 |
| Nivolumab | Adjuvant treatment of muscle invasive bladder cancer | 16.05 | \$1,657,990 |
| Olaparib | BRCA mutant Metastatic breast cancer second line treatment | 16.46 | \$644,871 |
| Nivolumab | Adjuvant treatment of oesophageal or gastroesophageal junctional adenocarcinoma | 17.20 | \$4,052,864 |
| Nivolumab | Metastatic Squamous cell carcinoma of oesophagus (second line) | 17.83 | \$1,417,085 |
| Nivolumab | Metastatic non-small cell lung cancer (2nd Line) | 17.83 | \$1,445,427 |
| Nivolumab | Metastatic renal cell carcinoma (2nd Line) | 18.16 | \$1,870,553 |
| Panitumumab | Metastatic Treatment Refractory Colorectal Cancer RAS wild type single agent use | 18.36 | \$2,261,307 |
| Pembrolizumab | Metastatic PD-L1 positive cervical cancer in combination with chemotherapy (1st Line) | 19.36 | \$2,708,543 |
| Pembrolizumab | Metastatic adenocarcinoma of oesophagus or gastroesophageal junction PD-L1 > 10% single agent (2nd Line) | 21.28 | \$571,608 |
| Cetuximab | Metastatic Left Sided Colorectal Cancer RAS wild type in combination with chemotherapy | 23.48 | \$13,065,327 |
| Pembrolizumab | Metastatic adenocarcinoma of oesophagus or gastroesophageal junction PD-L1 > 10% in combination with chemotherapy (1st Line) | 23.86 | \$2,515,075 |
| Vandatenib | Metastatic medullary carcinoma of thyroid | 26.54 | \$2,834,715 |
| Cetuximab | Unresectable squamous cell carcinoma of head and neck in combination with chemotherapy (1st Line) | 28.48 | \$7,335,075 |
| Pertuzumab Adjuvant Breast | Adjuvant treatment of HER2 positive breast cancer | 33.44 | \$23,741,644 |
| Nivolumab | Metastatic PD-L1 positive adenocarcinoma of oesophagus or gastroesophageal junction in combination with chemotherapy (first line) | 37.82 | \$2,579,095 |
| Tidal Cost per year | | | \$248,151,479 |

### Box 1

Cost-effective treatments not included in the World Health Organisation list of essential medicines

1. Abiraterone for hormone sensitive metastatic prostate cancer
2. Pomalidomide in the treatment of relapsed/refractory multiple myeloma
3. Topotecan in the second line treatment of advanced cervical cancer
4. Cabazitaxel in the treatment of metastatic castration resistant prostate cancer (post-docetaxel)
5. Fulvestrant in the first and second line treatment of endocrine sensitive metastatic breast cancer
6. Sunitinib in the first line treatment of metastatic renal cell carcinoma treatment of gastrointestinal stromal tumour (GIST)
7. Nab-paclitxael in the first line treatment of unresectable advanced pancreatic cancer
8. Bevacizumab for platinum refractory advanced epithelial ovarian cancer.
9. Trastuzumab for HER2 positive metastatic gastric cancer.
10. Adjuvant osimertinib in resected high risk EGFR mutation locoregional adenocarcinoma of lung
11. Adjuvant olaparib for germline BRCA mutation positive HER2 negative high risk early breast cancer
12. Sunitinib for first line treatment of metastatic renal cell cancer
13. Pazopanib for first line treatment of metastatic renal cell cancer
14. Sunitinib for second line treatment of unresectable gastrointestinal stromal tumours
15. Pazopanib for second line treatment of metastatic or unresectable soft tissue sarcoma
16. /Ribociclib for second line treatment of hormone sensitive metastatic breast cancer

## References

1. National Cancer Control Programme, Ministry of Health, Sri Lanka. Cancer Incidence Data - 2019. Available at https://www.nccp.health.gov.lk/storage/post/pdfs/Cancer%20Incidence%20Data%20Book-2019_compressed.pdf (Accessed 29th July 2022).

2. Joseph N, Gunasekera S, Ariyaratne Y, Choudhury A. Clinical Oncology in Sri Lanka: Embracing the Promise of the Future. Int J Radiat Oncol Biol Phys. 2019;105(3):466–70. doi:10.1016/j.radonc.2019.03.008

3. Gunasekera S, Seneviratne S, Wijeratne T, Booth CM. Delivery of cancer care in Sri Lanka.. J Cancer Policy. 2018;18:20–24. doi:10.1016/j.jcpo.2018.10.001

4. Ministry of Health Sri Lanka. Annual Health Bulletin 2019. pp 232. Available at: http://www.health.gov.lk/moh_final/english/public/elfinder/files/publications/AHB/2020/AHB%202019.pdf (Accessed 27th July 2022.)

5. Das M. Economic crisis in Sri Lanka causing cancer drug shortage. Lancet Oncol. 2022 Jun;23(6):710. doi: 10.1016/S1470-2045(22)00254-6

6. Medical Supplies Division, Ministry of Health, Sri Lanka. Standard Item Price List - 15.03.2022. Available at: https://www.msd.gov.lk/files/Pdf/price_list_2022.pdf (Accessed 27th July 2022)

7. US Dollar to Sri Lankan Rupee Spot Exchange Rates for 2021. Available at: https://www.exchangerates.org.uk/USD-LKR-spot-exchange-rates-history-2021.html x(Accessed 27th July 2022)

8. World Bank Data on Per Capita GDP - Sri Lanka. Available at https://data.worldbank.org/indicator/NY.GDP.PCAP.CD?locations=LK (Accessed 27th July 2022).

9. Bellido H, Olmos L, Román-Aso JA. Do political factors influence public health expenditures? Evidence pre- and post-great recession. Eur J Health Econ. 2019 Apr;20(3):455–474. doi: 10.1007/s10198-018-1010-2.

10. Chi YL, Blecher M, Chalkidou K, Culyer A, Claxton K et al. What next after GDP-based cost-effectiveness thresholds? Gates Open Res. 2020 Nov 30;4:176. doi: 10.12688/gatesopenres.13201.1

11. Edejer TT, Baltussen R, Tan-Torres T, Adam T, Acharya A et al (editors). Making choices in health: WHO guide to cost-effectiveness analysis. World Health Organisation; 2003.

12. Robinson LA, Hammitt JK, Chang AY, Resch S. Understanding and improving the one and three times GDP per capita cost-effectiveness thresholds. Health Policy Plan. 2017 Feb;32(1):141–145. doi: 10.1093/heapol/czw096.

13. Joseph N, Jayalth H, Balwardena J, Skandarajah T, Perera K. et al. Radical external beam radiotherapy in combination with brachytherapy for cervical cancer in Sri Lanka: Is treatment delayed treatment denied? JCO Global Oncology. 2020 Oct;6:1574–1581.

14. Rupasinghe T, Silva DC,Balawardena J, Perera K, Gunasekera D et al. Curative intent radiotherapy for squamous cell carcinoma of the head and neck in Sri Lanka: The impact of radiotherapy technique on survival. Clinical Oncology 2021; 33(12):765–772.

15. Hewage SA, Samaraweera S, Joseph N, Kularatna S, Gunawardena N. Presentation, Diagnosis and Treatment Delays in Breast Cancer Care and Their Associations in Sri Lanka, a Low-resourced Country. Clin Oncol (R Coll Radiol). 2022 Sep;34(9):598–607. doi: 10.1016/j.clon.2022.05.007.

16. Balawardena J, Skandarajah T, Rathnayake W, Joseph N. Breast Cancer Survival in Sri Lanka. JCO Global Oncology. April 2020. doi:10.1200/JGO.20.00003.

17. World Health Organisation Model List of Essential Medicines – 22nd List, 2021. Geneva: World Health Organisation; 2021 (WHO/MHP/HPS/EML/2021.02).

18. Wu YL, Tsuboi M, He J, John T, Grohe C et al. Osimertinib in Resected EGFR-Mutated Non-Small-Cell Lung Cancer. N Engl J Med. 2020 Oct 29;383(18):1711–1723. doi: 10.1056/NEJMoa2027071.

19. Tutt ANJ, Garber JE, Kaufman B, Viale G, Fumagalli D et al. Adjuvant Olaparib for Patients with BRCA1- or BRCA2-Mutated Breast Cancer. N Engl J Med. 2021 Jun 24;384(25):2394–2405. doi: 10.1056/NEJMoa2105215.

20. Szmulewitz RZ, Peer CJ, Ibraheem A, Martinez E, Kozloff MF et al. Prospective International Randomized Phase II Study of Low-Dose Abiraterone With Food Versus Standard Dose Abiraterone In Castration-Resistant Prostate Cancer. J Clin Oncol. 2018 May 10;36(14):1389-1395. doi: 10.1200/JCO.2017.76.4381.

21. Earl HM, Hiller L, Vallier AL, Loi S, McAdam K et al. 6 versus 12 months of adjuvant trastuzumab for HER2-positive early breast cancer (PERSEPHONE): 4-year disease-free survival results of a randomised phase 3 non-inferiority trial. Lancet. 2019 Jun 29;393(10191):2599–2612. doi: 10.1016/S0140-6736(19)30650-6.

22. National Comprehensive Cancer Network: Prostate Cancer, 2019. Available at: http://www.nccn.org/professionals/physician_gls/pdf/prostate.pdf

23. Morganti S, Bianchini G, Giordano A, Giuliano M, Curigliano G, Criscitiello C. How I treat HER2-positive early breast cancer: how long adjuvant trastuzumab is needed? ESMO Open. 2022 Apr;7(2):100428. doi: 10.1016/j.esmoop.2022.100428.

24. Li N, Zheng B, Cai HF, Yang J, Luo XF et al. Cost Effectiveness of Imatinib, Dasatinib, and Nilotinib as First-Line Treatment for Chronic-Phase Chronic Myeloid Leukemia in China. Clin Drug Investig. 2018 Jan;38(1):79–86. doi: 10.1007/s40261-017-0587-z.

25. Patel A, Goldstein DA, Tannock IF. Improving access to immunotherapy in low- and middle-income countries. Ann Oncol. 2022 Apr;33(4):360–361. doi: 10.1016/j.annonc.2022.01.003

