## Supplementary material for "Wisely Frugal: Ensuring sustainable funding for novel cancer therapeutics using cost-effectiveness thresholds in resource limited settings": Reference List

**List of References Used for Computation of Cost-effectiveness of each treatment**

| **Drug** | **Indication** | **Reference** |
| --- | --- | --- |
| Rituximab | Follicular lymphoma maintenance | J Clin Oncol. 2019 Nov 1;37(31):2815-2824. |
| Gefinib | Metastatic Non-small cell lung cancer 1st line EGFR mutation poitive | N Engl J Med 2010; 362:2380-2388 |
| Erlotinib | Metastatic Non-small cell lung cancer 1st line | Lancet Oncol 2011; 12: 735–42 |
| Everolimus | Advanced Renal Cell Carcinoma 2nd line (Vs placebo) | Cancer 2010 Sep 15;116(18):4256-65 |
| Pazopanib | Advanced Unresectable Sarcoma 2nd line | Lancet. 2012;379(9829):1879 |
| Palbociclib | Metastatic Hormone Sensitive Breast Cancer 2nd Line | N Engl J Med 2018; 379:1926-1936,N Engl J Med. 2015 Jul 16;373(3):209-19 |
| Lapatinib | Advanced HER2+ Breast Cancer Second Line treatment | N Engl J Med 2006; 355:2733-2743 |
| Palbociclib | Metastatic Hormone Sensitive Breast Cancer 1st Line | N Engl J Med 2016; 375:1925-1936 |
| Trabectidine | Advanced Unresectable Sarcoma 2nd line | J Clin Oncol. 2016;34(8):786 |
| topotecan | Advanced Platinum Refractory Epithelial Ovarian Cancer | J Clin Oncol. 1997; 15(6):2183-2193 |
| Ceritinib | Metastatic Metastatic Non-small cell lung cancer ALK+ 1st Line | Lancet. 2017;389(10072):917. |
| Bevacizumab | Metastatic Colorectal Cancer 1st line | J Clin Oncol. 2008 Apr 20;26(12):2013-9 |
| Crizotinib | Metastatic Metastatic Non-small cell lung cancer ALK+ 1st Line (vs chemo) | N Engl J Med 2013; 368:2385-2394 |
| Olaparib | Advanced Platinum Sensitive Epithelial Ovarian Cancer | J Clin Oncol. 2020;38(11):1164 |
| Nivolumab | Muscle Invasive Bladder Cancer Post Cystectomy Adjuvant Treatment | N Engl J Med 2021; 384:2102-2114 |
| Olaparib | Germline BRCA mutant and HER2-negative metastatic breast cancer | Ann Oncol. 2019;30(4):558 |
| Vandatenib | Advanced unresectable medullary carcinoma of thyroid | J Clin Oncol. 2012 Jan 10;30(2):134-41 |
| Nivolumab | Metastatic Melanoma | J Clin Oncol. 2020 Nov 20;38(33):3937-3946 |
| Pembrolizumab | Metastatic Melanoma | Lancet Oncol. 2019 Sep;20(9):1239-1251 |
| Sunitinib | Unresectable Gastrointestinal Stromal Tumour 2nd Line | Lancet. 2006;368(9544):1329, Clin Cancer Res. 2012 Jun 1; 18(11): 3170–3179. |
| Rituximab | Non-Hodgkin's Lympma1st line | N Engl J Med 2002; 346:235-242 Blood. 2014; 124(21):1752 and J Natl Cancer Inst. 2007 May 2;99(9):706-14 |
| Pembrolizumab | Metastatic Non-small cell lung cancer PD1> 50% Single agent 1st Line treatment | J Clin Oncol. 2019 Mar 1;37(7):537-546 |
| Olaparib | BRCA mutant Epithelial Ovarian Cancer First line Maintenance Treatment | Lancet Oncol 2021;22(12):1721-1731 |
| Pomalidamide | Multiple Myeloma 2nd line | Br J Haematol. 2015 Mar;168(6):820-3. |
| Trastuzumab | HER2+ Metastatic Breast Cancer | J Clin. Oncol. 2005 Jul 1;23(19):4265-74. |
| Nivolumab | Squamous cell carcinoma of the head and neck 2nd line | Oral Oncol . 2018 Jun;81:45-5 |
| Sunitinib | Advanced Renal Cell Carcinoma 1st Line | J Clin Oncol. 2009;27(22):3584-3590. |
| Pazopanib | Advanced Renal Cell Carcinoma 1st Line | N Engl J Med 2013;369:722-31. |
| Cetuximab | Treatment Refractory RAS wildtype Metastatic Colorectal Cancer (as monotherapy) | N Engl J Med 2008; 359:1757-1765 |
| Pembrolizumab | Unresectable PDL1+ Squamous cell carcinoma of head and neck 1st Line Single Agent Treatment | Lancet. 2019 Nov 23;394(10212):1915-1928 |
| Pembrolizumab | PDL1+ Metastatic Non-small cell lung cancer 2nd Line Single agent | Lancet 2016; 387: 1540–50 |
| Pembrolizumab | Metastatic Non-small cell lung cancer (Non-Squamous) 1st Line in combination with chemotherapy | J Clin Oncol 2020 May 10;38(14):1505-1517 |
| Panitumumab | Metastatic RAS wildtype colorectal cancer 1st line treatment in combination with FOLFOX | N Engl J Med 2013 Sep 12;369(11):1023-34. |
| Nivolumab | Adjuvant Treatment in Resected Oesophageal or Gastro-oesophageal Juntional Cancer | N Engl J Med 2021; 384:1191-1203 |
| Pembrolizumab | Metastatic Non-small cell lung cancer (Squmao)us 1st Line in combination with chemotherapy | J Thoracic Oncol. 2020 Oct;15(10):1657-1669. |
| Bortezomib | Multiple Myeloma (Trasnplant Inlegible) | Blood. 2011;118(21):476 |
| Pembrolizumab | Unresectable PDL1+ Squamous cell carcinoma of head and neck 1st Line treatment in combination with chemotherapy | Lancet. 2019 Nov 23;394(10212):1915-1928 |
| Nivolumab | Metastatic Non-small cell lung cancer 2nd line | J Clin Oncol. 2021 Mar 1;39(7):723-733 |
| Nivolumab | Advanced Renal Cell Carcinoma 2nd line | N Engl J Med 2015; 373:1803-1813 |
| Nivolumab | Metastatic Squamous Cell Carcinoma of Oesophagus 2nd line | Lancet Oncol. 2019 Nov;20(11):1506-1517. |
| Pertuzumab | Metastatic HER2+ Breast Cancer | N Engl J Med 2015; 372:724-734 |
| topotecan | Metastatic Cervical Cancer 2nd line | J Clin Oncol. 2005 Jul 20;23(21):4626-33 |
| Pembrolizumab | PD1+ Metastatic Cervical Cancer 1st line treatment in combination with chemotherapy | N Engl J Med 2021; 385:1856-1867 |
| Lenalidomide | Multiple Myeloma (Trasnplant Inlegible) | Blood 2018 Jan 18;131(3):301-310 |
| Bevacizumab | Platinum Refractory Epithelial Ovarian Cancer | J Clin Oncol. 2014;32(13):1302-8 |
| Panitumumab | Treatment Refractory RAS wildtype Metastatic Colorectal Cancer (as monotherapy) | Br J Cancer. 2016;115:1206-1214 |
| Pembrolizumab | Metastatic Triple Negative Breast Cancer PDL1 > 10% | N Engl J Med 2022; 387:217-226 |
| Bevacizumab | Metastatic Cervical Cancer | N Engl J Med 2014; 370:734-743 |
| Osimertinib | Metastatic EGFR mutant Non-small cell lung cancer 2nd line | Ann Oncol. 2020 Nov;31(11):1536-1544 |
| Ribociclib | Metastatic Hormone Sensitive Breast Cancer 2nd Line | N Engl J Med 2020; 382:514-524 |
| Nab-Paclitaxel | Advanced Pancreatic Cancer | N Engl J Med 2013; 369:1691-1703 |
| Pembrolizumab | Metastatic Oesophageal or Gastro-oesophageal Junctional Cancer PD1>10% 1st Line treatment in combination with chemotherapy | Lancet 2021; 398: 759–71 |
| Abiraterone 250 mg | Metastatic Hormone Sensitive Prostate Cancer | N Engl J Med 2017; 377: 338-51 and N Engl J Med. 2017;377:352-360 |
| Abiraterone 1000 mg | Metastatic Hormone Sensitive Prostate Cancer | Lancet Oncol. 2019;20(5):686-700 |
| Pembrolizumab | Metastatic Oesophageal or Gastro-oesophageal Junctional Cancer PD1>10% 1st Line treatment singe agent | J Clin Oncol. 2020 Dec 10;38(35):4138-4148 |
| Cetuximab | Unresectable Squamous cell carcinoma of Head and Neck 1st Line treatment in combination with chemotherapy | N Engl J Med 2008; 359:1116-1127 |
| Fulvestrant 500 mg | Metastatic Hormone Sensitive Breast Cancer 2nd Line | J Natl Cancer Inst. 2014 Jan;106(1):djt337 |
| Ribociclib | Metastatic Hormone Sensitive Breast Cancer 1st Line | N Engl J Med 2022; 386:942-950 |
| Decitabine | Acute Myeloid Leukaemia | J Clin Oncol. 2012. 30(21):2670-7 |
| Azacitadine | Acute Myeloid Leukaemia | Blood. 2015. 126(3):291-299. |
| Cetuximab | Metastatic RAS wildtype left sided colorectal cancer 1st line treatment in combination with chemotherapy | JAMA Oncol. 2017;3(2):194-20 |
| Enzalutamide | Metastatic Castration Resistant Prostate Cancer Post Docetaxel | N Engl J Med 2012; 367:1187-1197 |
| Olaparib | BRCA mutant epithelial ovarian cancer maintenance treatment post 2nd line chemotherapy | Lancet Oncol. 2021;22(5):620 |
| Sorafenib | Advanced hepatocellular carcionoma | N Engl J Med 2008; 359:378-390 |
| Fulvestrant 250 mg | Metastatic Hormone Sensitive Breast Cancer 1st Line | N Engl J Med 2019; 380:1226-1234 |
| Cabazitaxel | Metastatic Castration Resistant Prostate Cancer after docetaxel and novel anti-androgen | N Engl J Med 2019; 381:2506-2518 |
| Lenvatinib | Radioiodine Refractory Differentiated Thyroid Cancer | Eur J Cancer. 2021 Apr;147:51-57 |
| Regorafenib | Treatment Refractory Metastatic Colorectal Cancer | Lancet.2013;381(9863):303 |
| Abiraterone 250 mg | Metastatic Castration Resistant Prostate Cancer Post Docetaxel | N Engl J Med. 2011;3 64(21): 1995- 2005 |
| Abiraterone 1000 mg | Metastatic Castration Resistant Prostate Cancer Post Docetaxel | N Engl J Med. 2011;3 64(21): 1995- 2005 |
| Nivolumab | PDL1+ Metastatic gastro-eosphageal junctional or gasric cancer Metastatic 1st line | Lancet 2021 Jul 3;398(10294):27-40 |
| Trastuzumab | Metastatic HER2+ Gastric Cancer | Lancet 2010; 376: 687–97 |
| Bevacizumab | Metastatic Colorectal Cancer 2nd line | J Clin Oncol. 2007;25(12):1539-44 |
| Ribociclib | Metastatic Hormonse Sensitive Breast Cancer (Pre-menopausal) 1st Line | Clin Cancer Res. 2022 Mar 1;28(5):851-859 |
| Bevacizumab | Platinum Sensitive Relapsed Epithelial Ovarian Cancer | Lancet Oncol. 2017;18(6):779-791 |
| Bevacizumab | Metastatic Non-squmouas Non-small Cell Lung Cancer | N Engl J Med 2006; 355:2542-2550 |
| Bevacizumab | Metastatic Epithelial Ovary Cancer 1st line | Lancet Oncol 2015; 16: 928–36 |
| Osimertinib | Metastatic Non-small cell lung cancer 1st line | N Engl J Med 2020; 382:41-50 |
| Abiraterone 250 mg | Metastatic Castration Resistant Prostate Cancer Pre Docetaxel | Lancet Oncol. 2015 Feb;16(2):152-60. |
| Abiraterone 1000 mg | Metastatic Castration Resistant Prostate Cancer Pre Docetaxel | Lancet Oncol. 2015 Feb;16(2):152-60. |
| Nilotinib | Imatinib resistant Chronic Myeloid Leukaemia (in comparison to high dose imatinib) | Value Health. 2011 Dec;14(8):1057-67 |
| Enzalutamide | Metastatic Castration Resistant Prostate Cancer Pre Docetaxel | Eur Urol 2017;71(2):151-4 |
| Dasatinib | Imatinib resistant Chronic Myeloid Leukaemia (in comparison to high dose imatinib) | Value Health. 2011 Dec;14(8):1057-67 |
| Trastuzumab | Adjuvant Treatment of HER2+ Breast Cancer 6 months | Lancet. 2019;393(10191):2599-2612 |
| Trastuzumab | Adjuvant Treatment of HER2+ Breast Cancer 12 months | Lancet Oncol. 2021;22(8):1139-1150 |
| Goserelin | Adjuvant Treatment of Localised Prostate Cancer | J Clin Oncol 26: 2497–2504 |
| Osimertinib | Adjuvant treatmend of resected stage IB-III EGFR mutant adenocarcinoma of lung | N Engl J Med 2020; 383:1711-1723; Ann Oncol. 2022;33(Suppl_7):S1413-S1414 |
| Olaparib | BRCA mutant high risk resected early breast cancer | N Engl J Med 2021;384(25):2394 |
| Pembrolizumab | Stage III Resected Melanoma | Lancet Oncol 2021; 22: 643–54 |
| Nivolumab | Stage III Resected Melanoma | Eur J Cancer 2020 Jun;132:176-186 |
| Pembrolizumab | Adjuvant treatment of triple negative early breast cancer | N Engl J Med 2022; 386:556-567 |
| Pembrolizumab | Adjuvant treatment of resected Renal Cell Carcinoma | N Engl J Med 2021; 385:683-694 |
| Pertuzumab | Adjuvant treatment of HER2+ Breast Cancer | N Engl J Med 2017; 377:122-13 |
